# Exploring plasma cell motility and extracellular matrix protein biomarkers for primary progressive multiple sclerosis: A pilot study

**DOI:** 10.1101/2024.03.17.24304435

**Authors:** Elijah Lackey, Stephanie Reinke, Sheng Luo, Daniel Laskowitz, Christopher Eckstein, Simon G Gregory

## Abstract

Primary progressive multiple sclerosis is associated with neurodegeneration and chronic inflammation, and results in the accumulation of gradual disability. This pilot study investigated 92 plasma proteins using proximal extension assay to identify MS subtype-specific biomarkers with a focus on predicting primary progressive MS. We analyzed samples from 66 MS patients (22 relapsing-remitting, 22 secondary progressive, and 22 primary progressive) and 22 controls. ANOVA identified five proteins (ACAN, TMSB10, BST1, CLEC11A, MYOC) with p < 0.05 for differentiating phenotypes of MS, four of which have been previously implicated in MS pathophysiology. However, after correcting for multiple comparisons no individual proteins remained statistically significant. Logistic regression and support vector models using these 5 proteins for predicting primary progressive, in one-vs all-models, against other MS phenotypes and controls were of low accuracy (0.69 and 0.68, respectively). While not immediately translatable, these results lay the groundwork for future studies into MS progression biomarkers.

## Introduction

Multiple sclerosis (MS) is the most common autoimmune disease affecting the central nervous system. (1) MS causes both inflammatory relapses and a more insidious neurodegeneration referred to as progression. Recently, a paradigm shift has occurred in our understanding of disability accumulation in MS with the knowledge that progression contributes to disability accrual more than relapses.(2, 3) Multiple high-efficacy MS disease modifying therapies are available that effectively prevent relapses, but their effect on progression remains poorly understood. Because of this new focus on progression as a clinical target, there is greater need for disease-specific biomarkers that can monitor its development and track its severity.

Current blood biomarkers, such as neurofilament light chain and glial fibrillary acidic protein, can be increased by a wide variety of neurologic diseases.(4, 5) This lack of specificity has prevented their widespread clinical use for managing MS. Similarly, although some cerebrospinal fluid biomarkers such as SERPINA3 have shown promise, they are not practical for tracking disease activity longitudinally.(6) New disease and progression specific blood biomarkers will need to be discovered to translate effectively to the clinic.

In this pilot study, we targeted plasma proteins previously implicated in MS pathophysiology to identify specific markers of progression (Table 1). The panel we selected includes many proteins previously hypothesized to play a role in MS pathophysiology, neurodegeneration, or remyelination. We explored whether individual proteins or a group of proteins from this panel could distinguish between MS phenotypes and used selected proteins in a predictive model for primary progressive MS. Primary progressive MS was chosen as the phenotype of interest because these patients experience the greatest degree of progression.(3) A commercially available panel of proteins (Olink Development Target 96, Olink Waltham, MA) was selected that focuses on cell migration and extracellular matrix organization. such as Laminin A4, Aggrecan, and PARK7 (S1 Table).(7, 8, 9)

**Table 1.** Summary statistics after matching on age, sex, MS disease-modifying therapy, and race.

|  | Primary<br>Progressive | Secondary<br>Progressive | Relapsing<br>Remitting | Controls |
| --- | --- | --- | --- | --- |
| **Age, mean (SD)** | 57.4 (8.8) | 53.8 (9.9) | 49.9 (9.4) | 49.6 (9.7) |
| **Female Sex, n (%)** | 12 (54.5) | 17 (77.3) | 18 (81.8) | 15 (68.1) |
| **Most Common DMT** | Interferon<br>Beta-1a | Interferon<br>Beta-1a | Glatiramer<br>acetate | none |
| **Race, n (%)** |  |  |  |  |
| *White/Caucasian* | 18 (81.8) | 20 (90.9) | 18 (81.8) | 19 (86.4) |
| *Black/African American* | 4 (18.2) | 2 (9.1) | 4 (18.1) | 3 (13.6) |

## Materials and methods

Duke University IRB exemption was obtained (Pro00112531). Samples were obtained from adult patients (age >18) who participated in the MURDOCK (Measurement to Understand Reclassification of Disease Of Cabarrus and Kannapolis) study diagnosed with secondary progressive (SP), primary progressive (PP) or relapsing remitting (RR) MS. The MS cohort includes 25 PP patients, 88 SP patients, and 966 RR patients. Controls were derived from the greater MURDOCK study, which includes 12,526 patients. Samples were collected and stored (-80C) in a standardized process for the MURDOCK repository from 3/2009 to 8/2016. Clinical data was accessed August 4^th^, 2023. The authors did not have access to identifiable data for participants either before or after data collection. EDTA plasma samples were matched by age, sex, disease modifying therapy, and race (1:1:1:1). We excluded patients with recent radiographic relapse or treatment with steroids (within 30 days). MS diagnosis and relapses were self-reported by patients. Sample analysis was performed using Olink proximal extension assay, which provides a semi-quantitative measure of protein concentration called NPX (Normalized Protein eXpression) relative to controls. NPX units are the inverse of cyclic threshold (Ct) on a log2scale, which means the NPX value increases with greater protein concentrations. We ran 22 samples per group, each from a unique individual, for a total of 88 samples. We randomized samples across the 96 well plate. The kit was run with internal controls, negative controls, and sample controls.

The Olink platform has a standard quality control process involving four internal controls that were added to each sample to monitor assay performance and quality of individual samples. The sample plate was evaluated on the standard deviation to the internal controls. Samples that deviated less than 0.3 NPX from the median passed quality control. Primary statistical analysis was performed using one-way, balanced ANOVA on a per-protein basis. Bonferroni correction was applied to account for multiple comparisons. With 22 independent samples in each of the 4 groups, evaluating 92 proteins in each sample, we expected to detect an effect size of 0.55 with a power of 0.82 and an adjusted p-value of 0.0005. Proteins found to differentiate PP vs other subtypes of MS by ANOVA (p value < 0.05) were included in logistic regression and support vector machine models. We split pre-processed data into a training set (80%) and test set (20%). We performed 5-fold cross-validation to assess the performance of each model. Receiver operator curves were generated for the models and the area under the curve was calculated to measure performance of each model. Statistical analysis was done in Python (base 3.11.4) in a Jupyter notebook.

## Results

Samples were matched for age, sex, disease-modifying therapy (for MS patients), and race (Table 1). After randomization across the plate, samples were analyzed per the Olink platform protocol. All samples and assays met the platform’s quality control measures. ANOVA resulted in five proteins; aggrecan (ACAN), thymosin beta-10 (TMSB10), ADP-ribosyl cyclase (BST1), C-type lectin domain family 11 member A (CLEC11A), and myocilin (MYOC) with p-values less than 0.05. Although none of the proteins remaining statistically significant after Bonferroni correction (adjusted p value 0.0005), five proteins of interest were incorporated into logistic regression and support vector machine models comparing on an all-vs-one basis (PP vs all other groups including control). Logistic regression yielded an accuracy of 0.69 and the support vector machine an accuracy of 0.68 (Fig 1).

**Fig 1.**
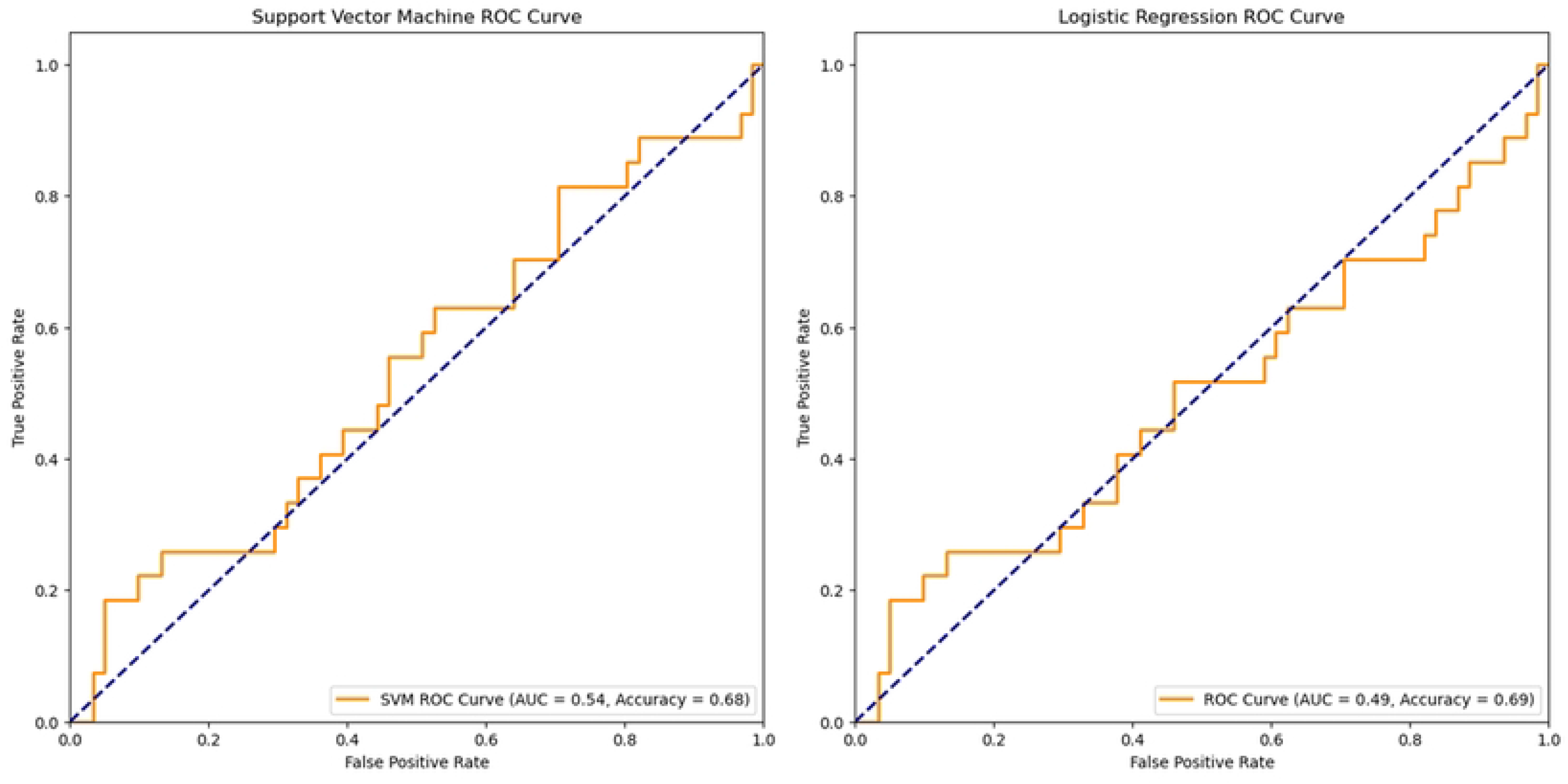
Receiver operating characteristic curves and accuracies for the support vector and logistic regression models.

## Discussion

As clinical priorities for the care of patients with MS shift toward a focus on progression, we need better biomarkers to quantify disease activity. We now know that progression plays a pivotal role in MS-related disability accrual.(3) This new perspective has sparked a need for more specific and readily accessible biomarkers to monitor and differentiate the extent of progression among MS patients. The choice of blood biomarkers in our study was deliberate, as they offer practicality and clinical utility. We refrained from revisiting well-established markers of neuronal injury and instead focused on a commercially available panel of proteins, Olink Development Target 96, that included multiple disease-specific proteins in order to improve the specificity of our results.

Our primary analysis yielded five proteins, namely aggrecan (ACAN), thymosin beta-10 (TMSB10), ADP-ribosyl cyclase (BST1), C-type lectin domain family 11 member A (CLEC11A), and myocilin (MYOC), with p-values below 0.05. Each of these were increased in PP vs other MS types except for myocilin (Fig 2).

**Fig 2.**
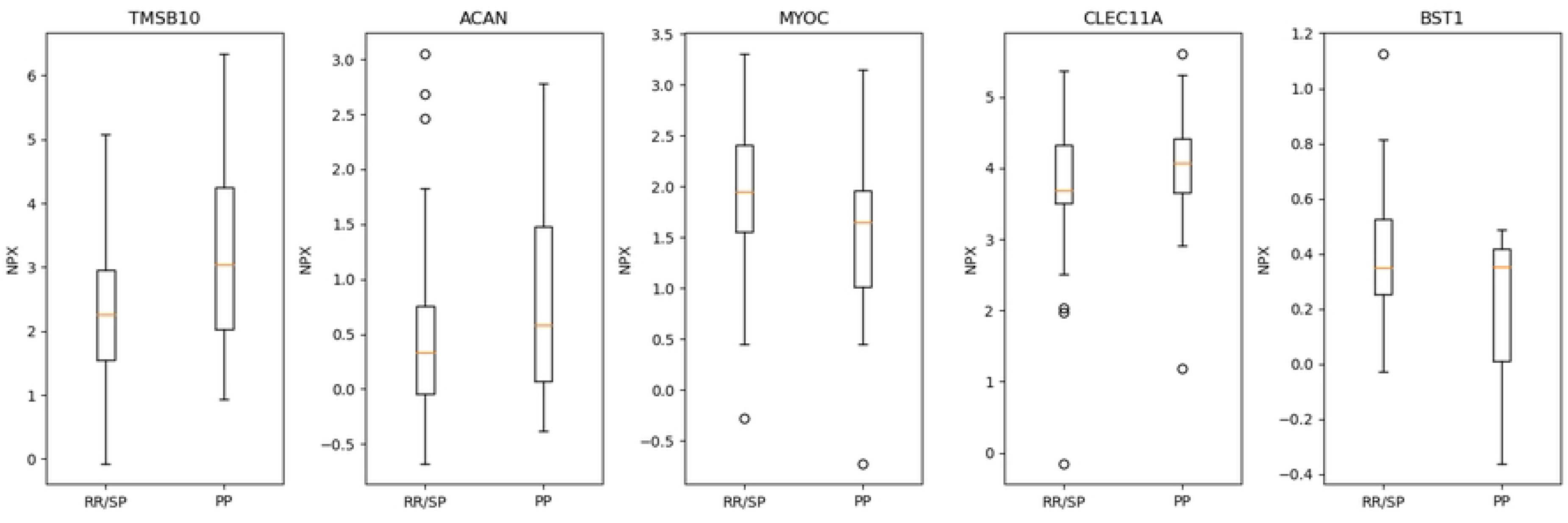
Concentrations of proteins by normalized protein expression units (NPX) found to distinguish the three MS subtypes (p <0.05) comparing RR grouped with SP vs PP. Each protein was increased in PP vs RR/SP except for myocilin.

However, after applying Bonferroni correction for multiple comparisons, none of these proteins remained statistically significant. To determine whether these 5 proteins might serve as biomarkers for predicting primary progressive MS we incorporated them into logistic regression and support vector machine models. These models attempted to distinguish PP from all three other groups (RR, SP, controls). Unsurprisingly, given our small sample size, these models had low accuracy (0.69 and 0.75 respectively).

It is important to note that four out of the five proteins; aggrecan (ACAN), ADP-ribosyl cyclase (BST1), thymosin beta-10 (TMSB10), and C-type lectin domain family 11 member A (CLEC11A), have previously been implicated in MS pathophysiology or in remyelination. ACAN, a proteoglycan, is known to be upregulated in MS lesions and may interfere with remyelination, while also playing a role in neurodegeneration. (8, 10, 11) BST1, thought to be involved with B cell maturation, may be upregulated when cognitive dysfunction is present in pediatric MS.(12) TMSB10, a member of the thymosin proteins, is part of a class of proteins that may regulate remyelination.(13) CLEC11A, a secreted sulfated glycoprotein acting on C-Type Lectin Receptors (CLRs), has been implicated in experimental autoimmune encephalomyelitis (EAE) pathogenesis.(14) Myocilin (MYOC), to our knowledge, has not been previously studied in MS disease pathophysiology. MYOC is a multimeric secreted glycoprotein that may affect leukocyte adhesion to endothelium.(15) Its specific role in the context of MS requires further exploration.

Our study has several limitations. Its reliance on self-reported MS diagnoses, based on outdated diagnostic criteria, is certainly a limitation. The relatively small sample size is also a constraint to be considered. Future studies with larger cohorts and samples from patients with more recent diagnoses, using the latest diagnostic criteria, are needed to advance our understanding of MS progression and to discover disease-specific biomarkers.

## Conclusions

This pilot study provides a preliminary exploration into plasma biomarkers for MS progression, with a specific focus on biomarkers predictive of primary progressive MS. Five proteins were identified; ACAN, BST1, TMSB10, CLEC11A, and MYOC that differentiate subtypes of MS with p <0.05. While none of the individual proteins achieved statistical significance after correction for multiple comparisons, these proteins show potential for further investigation. Importantly, myocilin (MYOC) is a secreted glycoprotein that has not previously been implicated in MS pathophysiology or investigated as a biomarker. The need for further research to validate and expand upon these findings remains pressing, particularly in the context of emerging treatments targeting MS progression.

## Data Availability

All data files, including Olink data and MS phenotypes correlating with samples, are available from the Open Science Framework database https://osf.io/7pj25/?view_only=49b942df41fe4e2e95bde4d4446d5488. This link is available for peer review and will be made public upon publication.

## Acknowlegments

This work was made possible through the use of the MURDOCK Study and Duke University’s CTSA grant (UL1TR002553) from the National Institutes of Health (NIH)’s National Center for Advancing Translational Sciences (NCATS).

## Supporting Information

**Supplemental Table 1.**
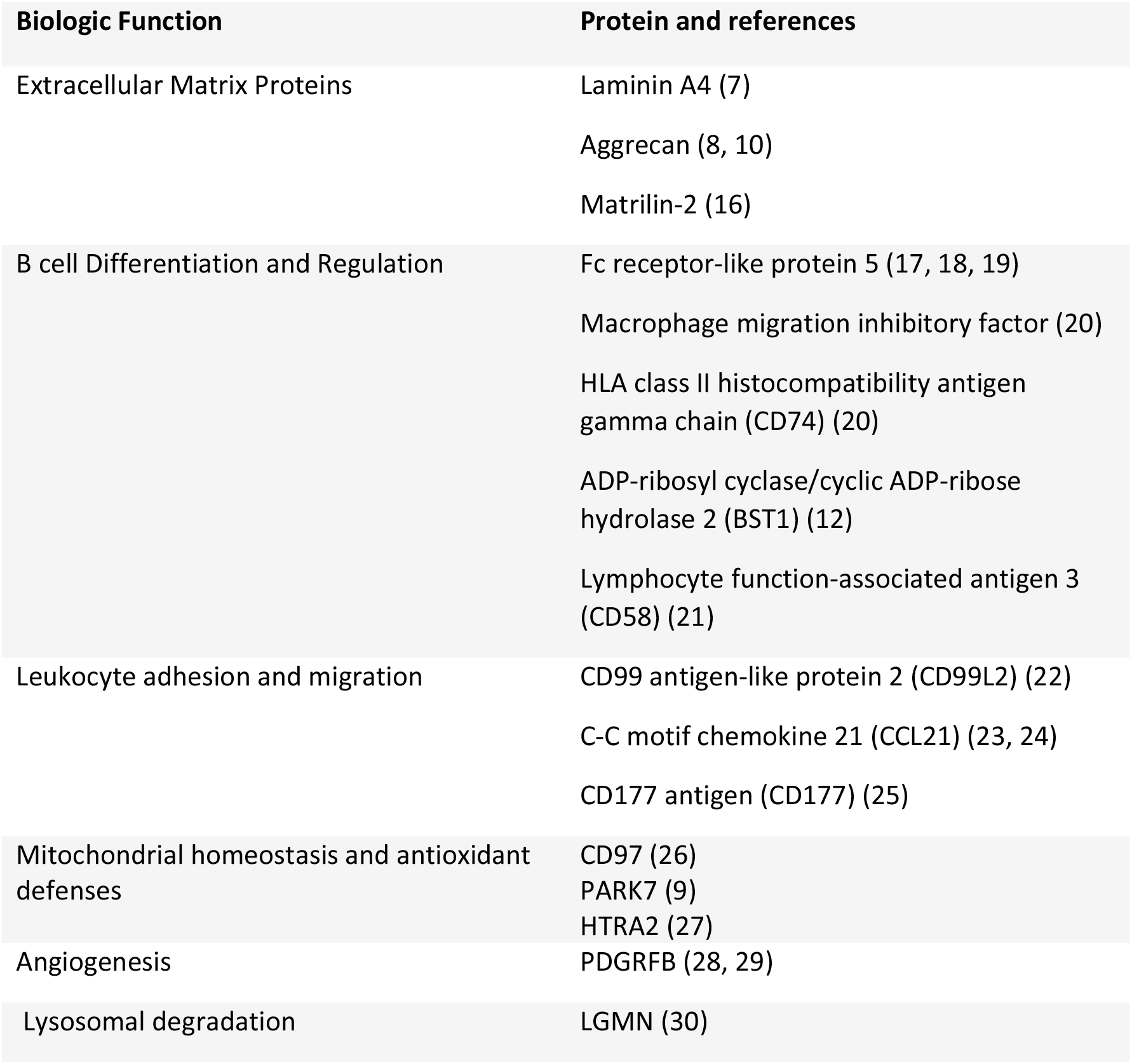
S1 Table. Proteins from the Olink Target 96 Development Panel Implicated in MS or EAE. Proteins in the Olink Target 96 Development Panel with prior evidence for involvement in experimental autoimmune encephalomyelitis (EAE) or multiple sclerosis (MS).

## Notes

### Competing Interest Statement

The authors have declared no competing interest.

### Funding Statement

The author(s) received no specific funding for this work.

### Author Declarations

Duke University Institutional IRB exemption was obtained (Pro00112531).

